# Browser-Based Video Supports Scalable Computer Vision Analysis for Open Surgical Skill Assessment

**DOI:** 10.64898/2026.11.09.26367680

**Authors:** Azad Khan, Soham Kulkarni, Shuo-Lun Lai, Xiaohan Wang, Idan Farber, Guillaume Kugener, Ankit Pal, Sofia Volpi, Orr Zohar, Chi-Sheng Chen, Serena Yeung-Levy, Gabriel Brat

## Abstract

**Purpose:** Objective assessment of open surgical skills remains limited by expert availability. Machine learning (ML)–based hand keypoint detection offers potential for automated, on-demand assessment, but performance in accessible browser-based recording environments (laptop-webcam video recorded in a web browser) is unknown. We hypothesized that browser-based recordings would achieve acceptable keypoint localization (percentage of correct keypoints [PCK] >70%).

**Methods:** Retrospective technical validation of prospectively collected video from 14 PGY-1 interns at one academic center (July to August 2024). We annotated 203 frames (101 browser-based, 1280×720/854×480; 102 overhead, 3840×2160/1920×1080), yielding 6,232 ground-truth keypoints. An internally developed pipeline and MediaPipe Hands were evaluated. The primary endpoint was PCK normalized to the index-proximal bone; bounding-box–normalized PCK, detection rate, and per-region accuracy were also evaluated.

**Results:** Primary PCK, normalized to a fixed anatomical hand scale, was 85.9% (95% CI, 80.3–91.0) for browser-based recordings and 81.5% (95% CI, 76.7–87.0) for overhead recordings. Secondary bounding-box–normalized PCK showed a similar pattern (91.6% vs 90.6%), and detection rates were high in both environments (94.9% vs 97.7%). In contrast, MediaPipe detection rates were only 5.3% for browser-based recordings and 15.4% for overhead recordings, resulting in substantially lower accuracy.

**Conclusion:** Hand keypoint detection performance was comparable between browser-based and overhead recordings using the internally developed pipeline. These findings suggest that browser-based video acquisition can provide data of sufficient quality for ML-based hand keypoint detection, offering a scalable pathway for surgical skill assessment without specialized recording hardware.

## Introduction

Surgical training has traditionally depended on subjective evaluations by experienced surgeons, with trainees acquiring skills through observation and hands-on practice [1]. However, expert availability for direct supervision is limited, creating challenges in training consistency and assessment reliability [1]. Established assessment instruments such as the Objective Structured Assessment of Technical Skills (OSATS) provide validated frameworks for evaluating surgical proficiency, but they require trained evaluators and are logistically constrained for frequent, formative use [2, 3]. Recent advances in machine learning (ML) and computer vision have enabled approaches to surgical skill assessment through automated video analysis [4–6].

Hand keypoint detection, the automated identification of anatomical landmarks on the hands, offers particular promise for assessing open surgical skills by enabling extraction of kinematic features (e.g., velocity) directly from video without specialized sensors [7]. Prior work has demonstrated that hand detection and tracking can distinguish between novice and experienced practitioners in open surgical tasks [8, 9], and that hand pose estimation can generate dexterity metrics with discriminative value [7]. However, these studies have largely relied on controlled laboratory or simulator setups with fixed camera viewpoints and overhead acquisition conditions. Whether machine learning models for hand keypoint detection can achieve comparable performance using laptop webcam videos acquired outside specialized recording environments remains unknown.

This distinction is critical for scalability. If accurate hand keypoint detection requires specialized recording hardware, deployment is limited to equipped simulation centers. If, however, standard webcam recordings provide sufficient signal quality, then ML-based assessment can be extended to any setting with a laptop, enabling distributed, on-demand training and data collection. We developed a browser-based platform that enables trainees to record open surgical tasks using a laptop webcam with a structured calibration workflow to standardize framing.

This study aimed to evaluate how recording modality affects hand keypoint detection performance by comparing results obtained from browser-based videos recorded via laptop webcams with those from overhead recordings, using manual annotations as the ground truth. We hypothesized that acceptable performance (percentage of correct keypoints [PCK] > 70%) could be achieved on browser-based recordings. This threshold was selected a priori based on the rationale that downstream kinematic feature extraction (e.g., velocity, path length) requires the majority of keypoints to be correctly localized to produce meaningful measurements. Secondary objectives included comparing performance against MediaPipe (an open-source hand-tracking solution) [10, 11], and characterizing performance variation across individual hand keypoints.

## Methods

### Study Design

This retrospective technical validation study evaluated the performance of an internally developed machine learning pipeline against manual ground-truth annotations for hand keypoint detection in surgical task videos. Videos were collected at Beth Israel Deaconess Medical Center (BIDMC) from July 2024 through August 2024 under a protocol approved by the BIDMC institutional review board (IRB) that authorizes video collection, annotation, and ML development for surgical education research. Oral informed consent was obtained from all participants.

### Participants and Video Sources

Surgical trainees enrolled in scheduled training sessions were eligible; no exclusion criteria were applied. Fourteen postgraduate year 1 (PGY-1) surgical interns participated. Participants uploaded a total of 321 videos through a custom browser-based recording platform designed to enable standardized recording of open surgical tasks using laptop webcams.

Two video datasets were used for the validation analysis:

#### Browser-based dataset

Videos were recorded by trainees using laptop webcams through the browser-based platform at resolutions of 1280 × 720 and 854 × 480. Before recording, participants completed a structured calibration workflow that guided laptop positioning, camera angle, task-field alignment, and hand placement to standardize framing and visibility of the task field. The laptop was positioned approximately 46 cm (18 inches) from the simulated tissue model, with the screen tilted toward the participant and the task field aligned to an on-screen guide. Recordings were then captured directly through the browser-based interface.

#### Overhead dataset

Overhead videos were recorded by interns from the same cohort, within an ongoing institutional research program conducted under the same IRB-approved protocol. These videos were recorded using a mounted GoPro camera positioned above the task field at a fixed distance, with consistent lighting and no camera movement. Recordings were captured at resolutions of 3840 × 2160 and 1920 × 1080 and included standardized open suturing tasks performed on a benchtop simulation setup. Both recording environments are shown in Figure 1.

**Fig. 1.**
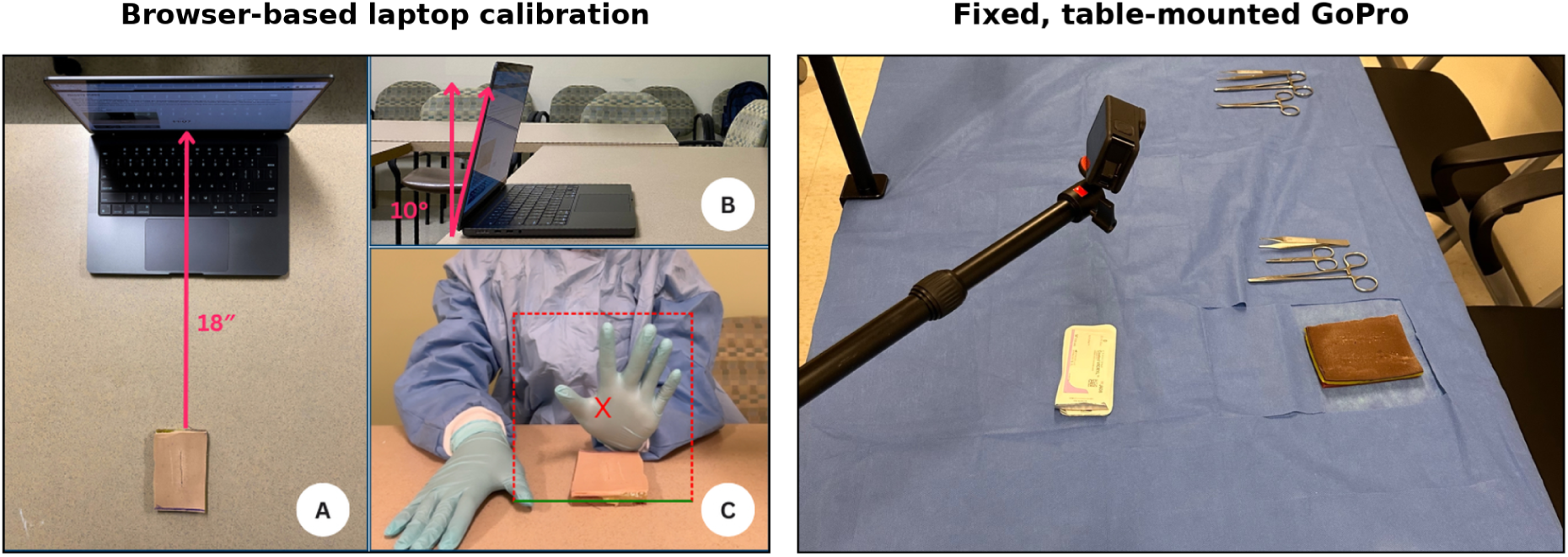
Recording environments. Left, structured laptop-webcam calibration workflow: (A) laptop positioned 46 cm (18 inches) from the front edge of the simulated tissue model; (B) screen tilted approximately 10° toward the participant; and (C) task field aligned with the on-screen guide and hand positioned within the calibration box. Right, fixed, table-mounted GoPro camera positioned above the benchtop suturing field

### Frame Sampling

Of the 321 uploaded browser-based videos, 29 were selected at random, yielding 101 annotated frames. Of the 720 available overhead videos, 22 were selected at random, yielding 102 annotated frames. All frames were sampled at 0.05 frames per second. In total, 203 frames were annotated, corresponding to 6,232 visible ground-truth keypoints (3,079 browser-based; 3,153 overhead).

### Internally Developed ML Pipeline

We used an internally developed two-stage pipeline built on two published models. Hands were first detected in each sampled frame with a Faster R-CNN hand detector [12], and 21 anatomical keypoints were then obtained for each detected hand with HaMeR [13], which reconstructs a three-dimensional hand model and projects its joints into the image.

The pipeline was applied to all sampled frames (0.05 frames per second) and output (x, y) coordinates for all 21 anatomical keypoints (standard finger-joint and palm landmarks) of each detected hand, regardless of keypoint visibility.

### Comparison: MediaPipe

MediaPipe Hands is an open-source hand landmark detection system developed by Google. In this study we used the MediaPipe Hand Landmarker (mediapipe version 0.10.21, Tasks application programming interface, full palm-detection and hand-landmark models) in image mode, predicting 21 anatomical keypoints for each detected hand, with a maximum of two hands per frame and the library’s default confidence thresholds of 0.5 for hand detection, hand presence, and tracking [10, 11]. MediaPipe was applied to the same sampled frames using the same preprocessing and evaluation steps as the internally developed pipeline. It was selected as a comparator because it is a widely used, publicly available hand-tracking solution.

Other available hand pose frameworks were considered; however, many do not distinguish left and right hands or are not designed for robust, direct inference on videos, making them less suitable as general-purpose comparators for this study. Because MediaPipe performance has been reported to depend on glove color [14], detection rate and PCK were additionally stratified by glove color, which was recorded for each video. Only two glove colors were present in the dataset, so this comparison is reported descriptively.

### Manual Ground-Truth Annotations

Ground-truth hand keypoints were manually annotated using the CVAT (Computer Vision Annotation Tool) platform [15] with a standardized 21-point hand landmark schema consistent with prior computer vision literature [16]. Annotators labeled all keypoints they judged to be visible and identifiable. To improve annotation efficiency, a model-assisted pre-annotation workflow was used. Model-generated keypoints were first uploaded into CVAT as an initial annotation layer, after which annotators manually reviewed, adjusted, and corrected the landmarks to create the final reference labels. Because pre-annotations were generated from model predictions, annotators were not blinded to the model output.

### Inter-Annotator Reliability

Two annotators independently labeled an overlapping set of 40 frames, including 20 browser-based frames and 20 overhead frames, to assess reliability of the ground truth. Localization agreement was quantified as the Euclidean distance between annotators, normalized separately by the index-proximal bone and by the hand bounding-box diagonal. Agreement on keypoint presence was quantified using Cohen’s κ.

### Definition of Keypoint Inclusion Criteria

Evaluation was restricted to visible keypoints that were identifiable and labeled by the annotators in the ground truth. Although the model output coordinates for all 21 keypoints per hand on every frame regardless of visibility, predictions corresponding to keypoints not labeled in the ground truth were excluded from analysis and were not counted as errors. This ensured that the model was neither penalized for predicting locations of non-visible keypoints nor credited for keypoints that could not be verified against ground truth.

## Analysis Methods

### Keypoint Localization Accuracy and Normalization

Keypoint localization accuracy was evaluated using percentage of correct keypoints (PCK). A predicted keypoint was counted as correct if its Euclidean distance from the corresponding annotated keypoint was within a specified fraction, α, of a reference hand scale. The primary reference scale was the index-proximal bone length, defined as the distance between the index-finger metacarpophalangeal and proximal-interphalangeal joints, corresponding to keypoints 5 and 6. This joint-to-joint anatomical reference provides a fixed hand-based scale that is independent of camera resolution, image size, and overall frame dimensions. This approach is consistent with established PCK-based pose-estimation evaluation, in which keypoint localization error is normalized by an object- or anatomy-specific reference distance rather than raw pixel distance [16].

The index-proximal bone was selected as the primary normalizer because its endpoints were the most consistently visible anatomical landmarks in the dataset and were present in approximately 97% of visible annotated keypoints. PCK was reported at α = 0.10, 0.20, and 0.30, with α = 0.30 used for the primary reported analysis. For each analysis, we also reported normalizer coverage, defined as the percentage of visible annotated keypoints for which the reference scale could be computed; keypoints were excluded from that analysis when the landmarks required for the scale were not visible.

As a secondary normalization strategy, PCK was recomputed using the diagonal of the ground-truth hand bounding box as the reference scale, evaluated at α = 0.05, 0.10, and 0.20. The bounding-box reference was defined for all annotated hands and was used as a robustness check to confirm that the primary findings were not driven by the small number of keypoints excluded when using the index-proximal bone reference.

### Performance Metrics

Percentage of correct keypoints quantified localization accuracy among visible annotated keypoints. Predicted keypoints were grouped into hands by the model’s left- or right-hand label (one predicted hand per label per frame). Within each frame, predicted hands were then optimally matched to annotated hands on the basis of the average distance between corresponding landmarks, without using the labels in the assignment (optimal assignment by the Hungarian algorithm). A pairing was accepted only if that average distance was no more than half the diagonal of the box enclosing the annotated hand’s landmarks; otherwise the annotated hand was counted as undetected. Landmarks within each matched pair were then compared one to one by anatomical position. Detection rate was reported separately as the proportion of visible ground-truth keypoints for which the model produced a matched prediction. Because each detected hand yielded all 21 keypoints, a keypoint was counted as undetected when its corresponding hand was not detected or not matched to the annotated hand. This allowed hand-detection failures to be distinguished from keypoint-localization errors.

Sensitivity analysis was performed using only keypoints for which all three candidate reference scales could be calculated: the index-proximal bone, palm length (wrist to the middle-finger metacarpophalangeal joint), and the bounding-box diagonal. This ensured that the normalizers were compared on the same set of keypoints. To maintain a comparable physical tolerance across scales, α was adjusted in inverse proportion to the median reference length.

### Per-Keypoint Analysis

To identify which anatomical landmarks were most and least accurately detected, PCK was calculated separately for each of the 21 hand keypoints, including the wrist, thumb joints, and the metacarpophalangeal (MCP), proximal interphalangeal (PIP), distal interphalangeal (DIP), and fingertip landmarks of the index, middle, ring, and little fingers. Per-keypoint metrics were computed separately for browser-based and overhead recordings.

### Comparison With MediaPipe

Model performance was compared with MediaPipe, an open-source hand-tracking solution [10, 11], using the same sampled frames, preprocessing approach, and evaluation pipeline. PCK and detection rate were computed for MediaPipe predictions and compared with the internally developed pipeline in both recording environments.

### Sample Size

Sample size was determined by annotation resource limitations. A total of 203 frames were annotated: 101 from 29 browser-based videos and 102 from 22 overhead videos, corresponding to 6,232 visible ground-truth keypoints (3,079 browser-based; 3,153 overhead). No formal sample size calculation was performed.

### Statistical Analysis

PCK was reported as the pooled proportion of correct keypoints across all visible keypoints for which the reference scale could be computed. To account for the clustering of keypoints within videos, 95% confidence intervals were obtained by a clustered bootstrap that resampled videos with replacement (10,000 resamples) and recomputed the pooled PCK. Recording environments were compared descriptively using point estimates and, for the primary endpoint, 95% confidence intervals; no formal hypothesis test was performed. Analyses were performed in Python 3.9 (NumPy 2.0.2, pandas 2.3.1, SciPy 1.13.1), with a fixed random seed for the bootstrap.

Claude (Anthropic) assisted in reviewing the analysis code; the code and results were verified by the authors who performed the analysis, and all authors take full responsibility for the content.

## Results

### Ground-Truth Quality

The median localization difference between annotators was 10.7% (IQR, 0.0–22.9) of the index-proximal bone length, which was within the primary PCK tolerance, and 2.5% (IQR, 0.0–5.4) of the hand bounding-box diagonal. Agreement on keypoint presence was substantial, with Cohen’s κ = 0.69, categorized as substantial agreement according to the conventional Landis and Koch interpretation [17].

### Dataset Characteristics

Table 1 summarizes the characteristics of both validation datasets.

**Table 1.** Characteristics of Validation Datasets.

| **Characteristic** | **Browser-Based** | **Overhead** |
|---|---|---|
| Videos sampled, No. | 29 | 22 |
| Frames annotated, No. | 101 | 102 |
| Resolution, pixels | 1280x720 + 854x480 | 3840x2160 + 1920x1080 |
| Visible ground-truth keypoints, No. | 3,079 | 3,153 |

### Model Performance

Using the internally developed pipeline, hand keypoint detection performance was comparable between browser-based and overhead recordings (Table 2). The primary anatomical reference, the index-proximal bone, was available for approximately 97% of visible annotated keypoints, including 96.6% of browser-based keypoints and 97.8% of overhead keypoints. Coverage for each candidate reference scale is reported in Online Resource 1, Table S3. At the primary threshold, PCK was 85.9% (95% CI, 80.3–91.0) for browser-based recordings and 81.5% (95% CI, 76.7–87.0) for overhead recordings. Both estimates exceeded the a priori acceptability threshold of PCK > 70%. The difference between recording environments was 4.4 percentage points (95% CI, -3.4 to 11.7).

**Table 2.** Keypoint Detection Performance by Recording Environment Using the Internally Developed Pipeline.

| **Performance Metric** | **Browser-Based** | **Overhead** |
|---|---|---|
| Index-proximal reference coverage, % | 96.6 | 97.8 |
| Primary PCK using index-proximal normalization (α = 0.30), % | 85.9 | 81.5 |
| Detection rate, % | 94.9 | 97.7 |
| Secondary PCK using bounding-box normalization (α = 0.10), % | 91.6 | 90.6 |

*Abbreviation: PCK, percentage of correct keypoints.*

This pattern was consistent under the secondary bounding-box normalization, where PCK was 91.6% for browser-based recordings and 90.6% for overhead recordings. Similar findings were observed in the sensitivity analysis comparing all three candidate normalizers on a common subset of keypoints, indicating that the result was not dependent on the choice of reference scale (Online Resource 1, Table S1). Detection rate was high in both environments, at 94.9% for browser-based recordings and 97.7% for overhead recordings, indicating that nearly all visible keypoints had matched model predictions. PCK across the full range of evaluated thresholds is shown for both normalizers in Figure 2 and tabulated in Online Resource 1, Table S2.

**Fig. 2.**
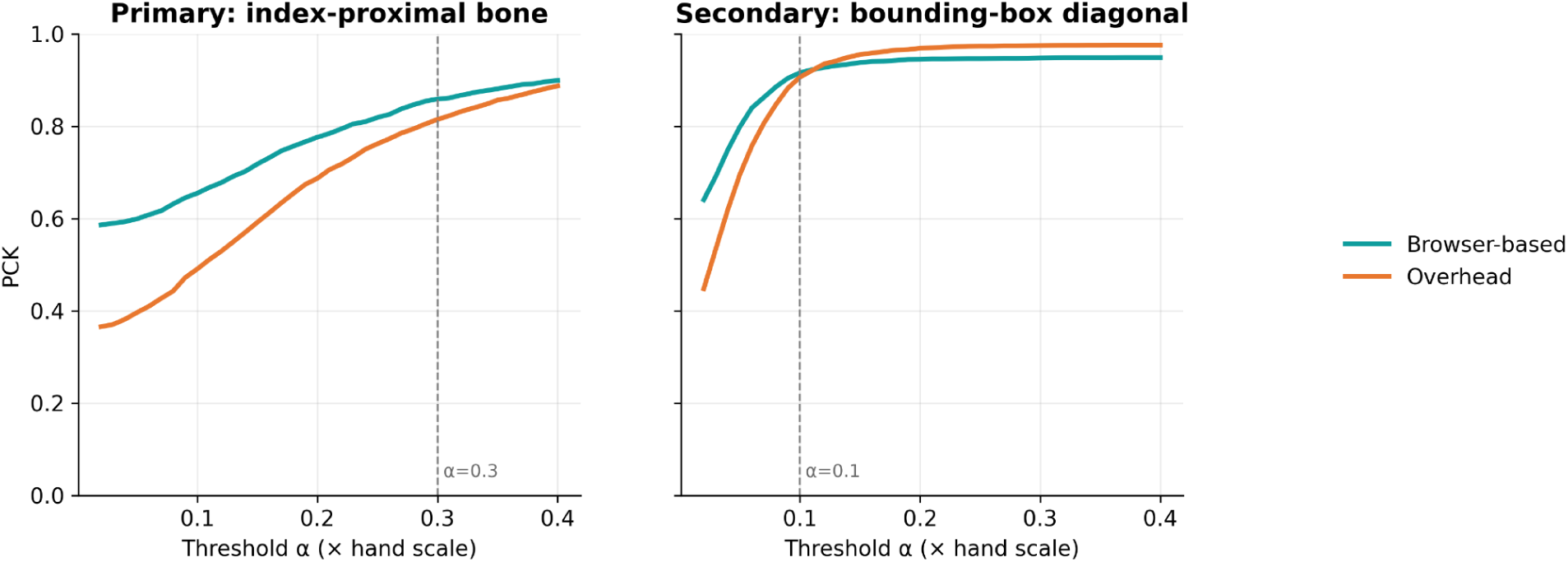
Percentage of Correct Keypoints Across Normalized Distance Thresholds. PCK for the internally developed pipeline across increasing normalized distance thresholds, α, comparing browser-based and overhead recordings. The primary analysis used the index-proximal bone as the reference scale, defined as the distance between the index-finger metacarpophalangeal and proximal-interphalangeal joints. The secondary analysis used the hand bounding-box diagonal as the reference scale

### Per-Keypoint Performance

In a secondary anatomical-region analysis using hand bounding-box normalization for full landmark coverage, localization accuracy was generally high across anatomical regions in both recording environments (Online Resource 1, Fig. S1). Per-finger PCK@0.10 was at least 87% in both browser-based and overhead recordings, indicating consistently high localization accuracy across the hand.

By joint level, the modest overhead deficit was concentrated at proximal landmarks, particularly the wrist (77% overhead vs 91% browser-based) and metacarpophalangeal joints (88% overhead vs 92% browser-based). In contrast, more distal landmarks, including the proximal-interphalangeal joints, distal-interphalangeal joints, and fingertips, were comparable between environments (91%–94%). No finger group demonstrated a marked performance drop in either environment.

The complete per-keypoint and joint-level breakdown across all 21 landmarks is provided in Online Resource 1, Tables S4 and S5.

### Comparison With MediaPipe

MediaPipe performed substantially worse than the internally developed pipeline in both recording environments (Table 3). MediaPipe detected only a minority of visible gloved surgical hands, with detection rates of 5.3% in browser-based recordings and 15.4% in overhead recordings. Because undetected keypoints were counted as localization failures, overall bounding-box–normalized PCK values were 4.2% for browser-based recordings and 11.3% for overhead recordings.

**Table 3.** Performance Comparison: Internally Developed Pipeline vs MediaPipe.

| **Metric** | **Browser-Based Internal** | **Browser-Based MediaPipe** | **Overhead Internal** | **Overhead MediaPipe** |
|---|---|---|---|---|
| Primary PCK using index-proximal normalization (α = 0.30), % | 85.9 | 3.5 | 81.5 | 9.1 |
| Detection rate, % | 94.9 | 5.3 | 97.7 | 15.4 |
| Secondary PCK using bounding-box normalization (α = 0.10), % | 91.6 | 4.2 | 90.6 | 11.3 |

*Abbreviation: PCK, percentage of correct keypoints.*

MediaPipe performance varied substantially by glove color. It detected no hands at all on blue gloves in either recording environment (0% detection across 31 videos), compared with 29.5% detection on purple gloves (20 videos). Performance of the internally developed pipeline was similar across the two glove colors evaluated, with detection rates of 96.7% versus 95.5% and PCK@0.10 of 90.8% versus 91.6% for blue and purple gloves respectively (Online Resource 1, Table S6).

## Discussion

This study demonstrates that hand keypoint detection performance was comparable between browser-based laptop recordings and overhead recordings when evaluated using normalized localization metrics. Using the internally developed pipeline, keypoint localization accuracy was similar across recording environments, with closely overlapping confidence intervals between modalities. The model also achieved high detection rates in both settings and substantially outperformed MediaPipe, which detected only a minority of gloved surgical hands.

### Implications for Scalable Surgical Skill Assessment

For the internally developed pipeline, the overhead setup, which combined higher spatial resolution with fixed-camera mounting, did not improve keypoint localization compared with browser-based recordings. Because these acquisition factors differed together, the findings reflect the overall recording setup rather than the effect of spatial resolution alone.

From a practical perspective, specialized recording hardware does not appear to be a prerequisite for ML-based hand keypoint detection in this setting. Because a laptop webcam and a browser-based calibration workflow are available wherever trainees already work, this could reduce a major hardware barrier to collecting surgical video at scale. Standard laptop webcams, when combined with structured calibration procedures, may provide sufficient video quality for scalable surgical skill assessment and distributed data collection. The calibration workflow built into the browser-based platform, which guides framing and hand scale, may help offset the lower resolution of webcam video. Because the approach requires only a laptop and a browser, it may also extend objective skill assessment to resource-constrained settings where dedicated recording hardware is unavailable. Beyond the assessment of individual trainees, this has implications for how open surgical video datasets are built. In open surgery, progress in computer vision has been limited by a lack of large annotated video datasets [18], and collecting such video has typically required dedicated cameras and fixed installations. A browser-based workflow shifts collection onto hardware trainees already use: in this study, 14 interns contributed 321 recordings over two months. Data gathered this way could support model development, curriculum evaluation, and longitudinal performance feedback across multiple institutions.

### Comparison With MediaPipe

MediaPipe demonstrated substantially lower performance than the internally developed pipeline across both datasets. Its poor performance primarily reflected failure to detect gloved surgical hands, suggesting that the model did not generalize well to this surgical video setting.

According to its original description, MediaPipe Hands was trained on a combination of in-the-wild real-world images, an in-house gesture dataset, and synthetic hand renderings, primarily representing bare hands in general-purpose environments [10]. The published training datasets do not include surgical scenes, gloved hands, or instrument interactions. The poor performance observed in our study may therefore reflect limited representation of surgical-domain visual characteristics in its training data.

Prior surgical-domain studies have reported that MediaPipe performance decreases in the presence of surgical gloves and varies substantially by glove color, with particularly poor localization for blue and green gloves [14]. Similarly, Kiefer et al. reported that MediaPipe Hands failed to detect keypoints on medical-gloved hands, whereas a domain-specific DeepLabCut model successfully tracked gloved-hand landmarks [19]. In our own data this pattern was reproduced directly: MediaPipe failed entirely on blue gloves in both recording environments, whereas performance of the internally developed pipeline was similar across the two glove colors evaluated. These findings suggest that lightweight general-purpose hand trackers may not generalize to gloved surgical hands.

Future work should evaluate whether keypoint-derived kinematic features correlate with validated measures of surgical proficiency and determine the robustness of these findings across multiple institutions and recording setups.

## Limitations

Manual annotation served as the ground truth. Although such labeling is inherently subjective, inter-annotator agreement was substantial. However, annotation was model-assisted: model predictions served as initial labels, which annotators then reviewed and corrected. Absolute accuracy may therefore be overestimated, and because both annotators began from the same initial labels, inter-annotator agreement reflects the reproducibility of the ground truth rather than an independent estimate of human annotation accuracy. As the same annotation protocol was applied to both recording environments, this bias is expected to affect absolute accuracy more than the comparison between environments.

A second limitation is that PCK estimates depend on the choice of hand-scale normalizer. We used the index-proximal bone as the primary anatomical reference and also reported results using the hand bounding-box diagonal as a secondary measure with full keypoint coverage. Sensitivity analysis on the subset of keypoints for which all three candidate scales were available showed similar findings across normalization approaches. However, the choice of normalizer and the α threshold was made during the analysis rather than prespecified. Although this approach reduces the influence of normalization strategy, it does not eliminate it entirely.

The study was conducted at a single academic center. Participants were enrolled by convenience from scheduled training sessions, although videos were selected at random from those available, which may limit generalizability to other institutions, recording setups, or surgical tasks. The sample size was determined by annotation resources rather than a formal power calculation, and the study evaluated only suturing tasks, which may not generalize to other open surgical procedures. Finally, the browser-based and overhead environments differ in several bundled factors: spatial resolution, camera mounting and viewpoint, and framing. Therefore, the observed comparability reflects the overall acquisition setting rather than the effect of resolution alone.

## Conclusions

Hand keypoint detection performance was comparable between browser-based laptop recordings and overhead recordings when evaluated using a fixed anatomical normalizer, with detection reported separately from localization. Performance in both environments exceeded the a priori acceptability threshold of 70%. In the secondary bounding-box–normalized analysis, the internally developed pipeline demonstrated high localization accuracy across anatomical regions in both recording environments, with no landmark group showing a marked performance failure. These findings indicate that accessible browser-based video can provide data of sufficient quality for ML-based hand keypoint detection, supporting a scalable approach to surgical skill assessment and model development. In contrast, a lightweight hand-tracking model developed for bare hands did not generalize to gloved surgical hands.

## Supporting information

Online Resource 1: Tables S1-S6 and Fig. S1

## Statements and Declarations

### Ethics approval

This study was approved by the Beth Israel Deaconess Medical Center Institutional Review Board and conducted in accordance with the Declaration of Helsinki.

### Consent to participate

Oral informed consent was obtained from all participants.

### Consent to publish

Not applicable; no identifiable images of participants are included.

### Data availability

The data are restricted under the IRB-approved protocol and are not publicly available. The analysis code and pipeline are available from the corresponding author on reasonable request.

