## Supplementary material for "Browser-Based Video Supports Scalable Computer Vision Analysis for Open Surgical Skill Assessment": Online Resource 1: Tables S1-S6 and Fig. S1


1. Normalizer sensitivity analysis
2. PCK across all evaluated thresholds
3. Normalizer coverage
4. Per-keypoint performance (all 21 landmarks)
5. Joint-level and finger-group performance
6. Glove color analysis
7. Anatomical Group-Level Keypoint Detection Accuracy by Recording Environment


**Table S1. Normalizer sensitivity analysis**

PCK computed under three hand-scale normalizers on the subset of keypoints for which all three scales were defined, with each threshold calibrated to a common physical tolerance (10% of the hand bounding-box diagonal, equivalent to α = 0.44 and 0.39 for the index-proximal bone and α = 0.25 and 0.21 for palm length, in browser-based and overhead recordings, respectively). Estimates are concordant, indicating the primary result does not depend on the choice of reference scale. Analysis restricted to the subset of keypoints for which all three reference scales could be computed (browser-based n = 2,757 of 3,079; overhead n = 2,727 of 3,153), so that the normalizers are compared on identical keypoints. The subset is limited by palm-length coverage, which requires both the wrist and middle-finger MCP to be visible.

| **Recording environment** | **Keypoints (n)** | **Index-proximal, %** | **Palm-length, %** | **Bounding box, %** |
|---|---|---|---|---|
| Browser-based | 2,757 | 91.3 | 90.4 | 92.3 |
| Overhead | 2,727 | 89.0 | 85.9 | 90.8 |

**Table S2. PCK across all evaluated thresholds**

Percentage of correct keypoints at every threshold reported in the Methods, for both normalizers, the internal pipeline and MediaPipe, and both recording environments. Bold values are the internal pipeline values reported in the main text (primary: index-proximal α=0.30; secondary: bounding box α=0.10).

| **Model/Pipeline** | **Environment** | **Idx-prox α=0.10** | **Idx-prox α=0.20** | **Idx-prox α=0.30** | **Bbox α=0.05** | **Bbox α=0.10** | **Bbox α=0.20** |
|---|---|---|---|---|---|---|---|
| Internal pipeline | Browser-based | 65.5 | 77.6 | **85.9** | 79.9 | **91.6** | 94.5 |
| Internal pipeline | Overhead | 49.1 | 68.7 | **81.5** | 69.5 | **90.6** | 96.9 |
| MediaPipe | Browser-based | 0.9 | 2.3 | 3.5 | 2.5 | 4.2 | 4.9 |
| MediaPipe | Overhead | 2.3 | 6.0 | 9.1 | 6.3 | 11.3 | 14.9 |

Abbreviations: Bbox, bounding box; Idx-prox, index-proximal.

**Table S3. Normalizer coverage**

Percentage of visible annotated keypoints for which each hand-scale reference could be computed. The index-proximal bone was selected as the primary normalizer because it had the highest coverage of the anatomical scales; the bounding box (100% coverage) is used as the secondary reference.

| **Recording environment** | **Index-proximal, %** | **Palm-length, %** | **Bounding box, %** |
|---|---|---|---|
| Browser-based | 96.6 | 91.9 | 100.0 |
| Overhead | 97.8 | 88.6 | 100.0 |

**Table S4. Per-keypoint performance (all 21 landmarks)**

PCK@0.10 and detection rate for each anatomical landmark, using bounding-box normalization (full landmark coverage).

| **Landmark** | **Browser-based PCK@0.10, %** | **Browser-based detection, %** | **Overhead PCK@0.10, %** | **Overhead detection, %** |
|---|---|---|---|---|
| Wrist | 91.0 | 94.2 | 76.7 | 97.2 |
| Thumb CMC | 92.6 | 93.9 | 84.0 | 97.0 |
| Thumb MCP | 87.3 | 94.0 | 82.5 | 96.8 |
| Thumb IP | 86.8 | 94.6 | 90.6 | 96.6 |

| | | | | |
|---|---|---|---|---|
| Thumb Tip | 91.9 | 96.7 | 91.8 | 96.8 |
| Index MCP | 94.4 | 94.9 | 93.0 | 97.5 |
| Index PIP | 94.4 | 94.9 | 97.0 | 97.5 |
| Index DIP | 93.7 | 94.7 | 94.8 | 97.4 |
| Index Tip | 94.8 | 94.8 | 96.0 | 98.3 |
| Middle MCP | 94.4 | 94.9 | 83.7 | 97.4 |
| Middle PIP | 92.3 | 94.9 | 94.1 | 97.8 |
| Middle DIP | 92.6 | 94.9 | 92.3 | 98.7 |
| Middle Tip | 92.5 | 94.6 | 93.0 | 98.2 |
| Ring MCP | 91.1 | 94.8 | 91.7 | 98.2 |
| Ring PIP | 90.4 | 94.9 | 93.0 | 97.5 |
| Ring DIP | 89.0 | 95.6 | 93.5 | 98.1 |
| Ring Tip | 85.7 | 93.7 | 89.0 | 97.3 |
| Pinky MCP | 91.6 | 96.4 | 92.5 | 98.0 |
| Pinky PIP | 90.1 | 95.4 | 94.9 | 98.5 |
| Pinky DIP | 88.9 | 95.1 | 90.0 | 98.9 |

| | | | | |
|---|---|---|---|---|
| Pinky Tip | 82.1 | 94.6 | 88.6 | 100.0 |

Abbreviations: CMC, carpometacarpal; DIP, distal interphalangeal; IP, interphalangeal; MCP, metacarpophalangeal; PCK, percentage of correct keypoints; PIP, proximal interphalangeal.

**Table S5. Joint-level and finger-group performance**

PCK@0.10 (bounding-box normalized) pooled by joint level (proximal to distal) and by finger group.

| **Grouping** | **Region** | **Browser-based PCK@0.10, %** | **Overhead PCK@0.10, %** |
|---|---|---|---|
| Joint level | Wrist | 91.0 | 76.7 |
| Joint level | MCP | 92.2 | 87.9 |
| Joint level | PIP | 91.2 | 94.1 |
| Joint level | DIP | 91.8 | 93.0 |
| Joint level | Tip | 91.1 | 92.8 |
| Finger group | Thumb | 89.7 | 87.1 |
| Finger group | Index | 94.3 | 95.2 |
| Finger group | Middle | 93.1 | 90.3 |
| Finger group | Ring | 89.9 | 92.1 |
| Finger group | Pinky | 89.5 | 92.3 |

Abbreviations: DIP, distal interphalangeal; MCP, metacarpophalangeal; PCK, percentage of correct keypoints; PIP, proximal interphalangeal.

**Table S6. Glove color analysis**

Detection rate and PCK@0.10 (bounding-box normalized) stratified by glove color, within each recording environment and pooled. MediaPipe detected no hands wearing blue gloves in either environment, indicating color-dependent failure; the internally developed pipeline performed similarly across the two glove colors.

| Recording environment | Model/Pipeline | Glove color | Videos (n) | Detection, % | PCK@0.10, % |
|---|---|---|---|---|---|
| Browser-based | Internal pipeline | Blue | 20 | 94.9 | 91.5 |
| Browser-based | Internal pipeline | Purple | 9 | 95.0 | 91.8 |
| Browser-based | MediaPipe | Blue | 20 | 0.0 | 0.0 |
| Browser-based | MediaPipe | Purple | 9 | 19.6 | 15.6 |
| Overhead | Internal pipeline | Blue | 11 | 99.1 | 89.9 |

| | | | | | |
|---|---|---|---|---|---|
| Overhead | Internal pipeline | Purple | 11 | 95.9 | 91.6 |
| Overhead | MediaPipe | Blue | 11 | 0.0 | 0.0 |
| Overhead | MediaPipe | Purple | 11 | 35.3 | 25.9 |
| Both | Internal pipeline | Blue | 31 | 96.7 | 90.8 |
| Both | Internal pipeline | Purple | 20 | 95.5 | 91.6 |
| Both | MediaPipe | Blue | 31 | 0.0 | 0.0 |
| Both | MediaPipe | Purple | 20 | 29.5 | 22.0 |

**Fig. S1. Anatomical Group-Level Keypoint Detection Accuracy by Recording Environment**

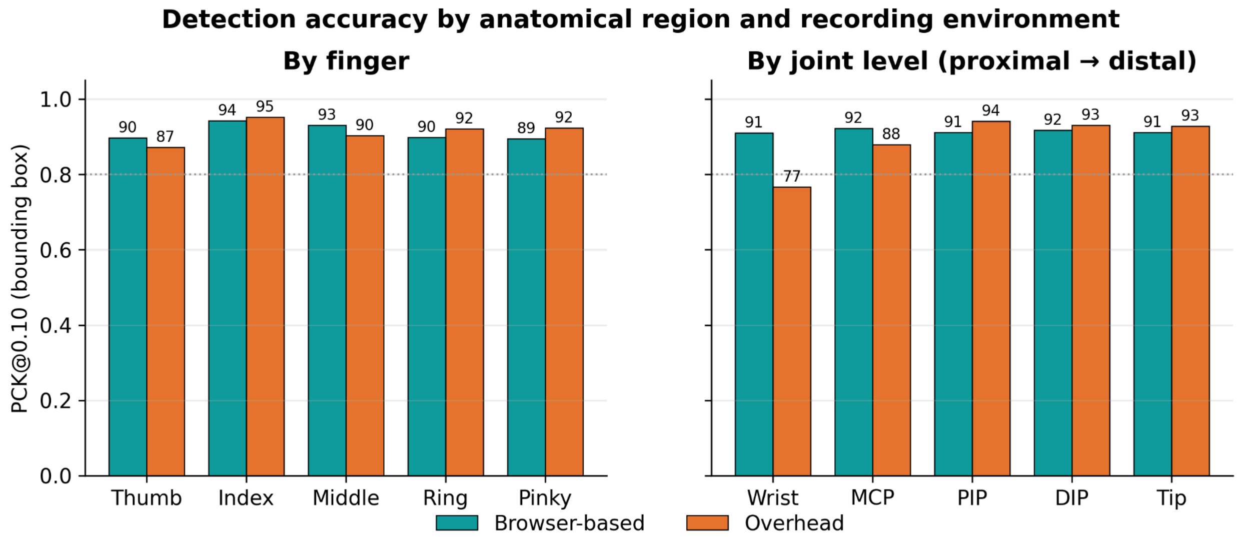

*Percentage of correct keypoints (PCK@0.10) by anatomical region: (left) by finger and (right) by joint level (proximal to distal), comparing browser-based and overhead recordings. PCK was calculated using hand bounding-box normalization to provide full landmark coverage across the hand.*
